# Comparison of Cross-Table and Lauenstein Lateral Views for Tip Apex Distance Measurement in Trochanteric Fracture Fixation

**DOI:** 10.1101/2025.10.05.25337377

**Authors:** Daiki Saito, Naoya Hoshikawa, Takuya Uematsu, Makoto Hirao

**Author notes:** Corresponding author: Daiki Saito, MD, Department of Orthopaedic Surgery, Nippon Medical School, 1-1-5 Sendagi, Bunkyo-ku, Tokyo, Japan.

## Abstract

**Background:** Tip apex distance (TAD) is a commonly used radiographic index to evaluate the risk of mechanical failure following internal fixation of trochanteric femoral fractures. While the cross-table lateral view (C view) is typically used to assess TAD, Lauenstein’s I view (L view), which approximates the true lateral view, may also allow for reliable measurement. However, the consistency of TAD measurements across different lateral projections has not been fully investigated.

**Methods:** We conducted a retrospective cohort study of patients who underwent internal fixation with a short femoral nail for trochanteric fractures between July 2016 and January 2020. Patients who had both C view and L view radiographs postoperatively were included. TAD was measured on each view and compared using paired t-tests. Pearson’s correlation coefficient was calculated, and agreement was evaluated using Bland–Altman analysis.

**Results:** A total of 31 patients were included. The mean TAD was 8.55 mm (SD, 2.40; 95% CI, 7.62–9.48) in the C view and 9.63 mm (SD, 3.34; 95% CI, 8.41–10.84) in the L view, with a statistically significant mean difference of 1.06 mm (p = 0.027; 95% CI, 0.13–2.03). A moderate positive correlation was observed between the two views (r = 0.637, p < 0.001). Bland–Altman analysis showed good agreement between the two measurements.

**Conclusions:** Although a statistically significant difference in TAD measurements was observed between C view and L view, the correlation and agreement were relatively high. These findings suggest that L view may serve as a practical alternative when obtaining the C view is not feasible. Further studies are warranted to confirm these results and evaluate their clinical implications.

## Introduction

Tip apex distance (TAD) is widely used as a radiographic index to evaluate the risk of cut-out following internal fixation of trochanteric femoral fractures. The concept, originally introduced by Baumgaertner et al., combines measurements from anteroposterior and lateral radiographs, with a threshold of <25 mm being recommended to minimize the risk of failure.(1) TAD represents the sum of the distances from the tip of the lag screw to the apex of the femoral head measured on anteroposterior (AP) and lateral radiographs, corrected for magnification. It has been shown to be a strong predictor of lag screw cut-out, a common complication following internal fixation

However, the accuracy of TAD is inherently dependent on radiographic technique, particularly the lateral view. Geller et al. and Nikoloski et al. have demonstrated that deviations in radiographic angles—such as malrotation or suboptimal lateral projection—can lead to significant discrepancies in TAD values, potentially affecting clinical decision-making.(2)(3) According to the Japanese Guidelines for the Treatment of Femoral Neck and Trochanteric Fractures (2021), TAD should ideally be measured using the cross-table lateral view (C view).(4)

Obtaining an optimal cross-table lateral view can be technically challenging in elderly patients or those with limited mobility due to pain, postoperative condition, or body habitus. In contrast, Lauenstein’s I view (L view) can be performed with the hip flexed and externally rotated, which often results in more consistent imaging conditions. Herngren et al. reported excellent inter- and intraobserver reliability for measuring slip angle on L view (ICC 0.98– 0.99), and Terjesen et al. demonstrated substantial reliability (ICC ≥ 0.75) for radiographic parameters using lateral hip radiographs (5)(6). These findings suggest that L view may provide more consistent imaging conditions than the cross-table lateral view in certain clinical settings, which could help improve the reliability of postoperative radiographic assessment.

The true lateral view, which corrects for femoral neck anteversion, is considered more anatomically accurate for evaluating the proximal femur. L view, which is commonly used for evaluating the proximal femur, provides a projection that closely resembles the true lateral orientation. However, few studies have compared the consistency of tip apex distance (TAD) measurements between L view and the standard cross-table lateral view. We therefore aimed to assess whether TAD measurements obtained from L view are comparable to those from C view.

This study aimed to assess the effect of radiographic technique on TAD by comparing measurements obtained from C view and L view, and to explore the potential interchangeability of these projections in postoperative evaluation.

## Materials and Methods

This retrospective cohort study included patients who underwent internal fixation with a short femoral nail for trochanteric femoral fractures at Nippon Medical School Hospital between July 2016 and January 2020. Patients with both postoperative C view and L view radiographs were eligible.

TAD was measured using standard digital radiographic techniques for each view. When calculating TAD, the distance from the tip of the lag screw to the apex of the femoral head on lateral radiographs was measured along the axis of the femoral neck, as shown in Figure 1. Paired t-tests were used to compare mean TAD values between C view and L view. Pearson’s correlation coefficient was calculated to assess the linear relationship, and Bland–Altman analysis was used to evaluate the agreement between the two methods.

**Figure 1.**
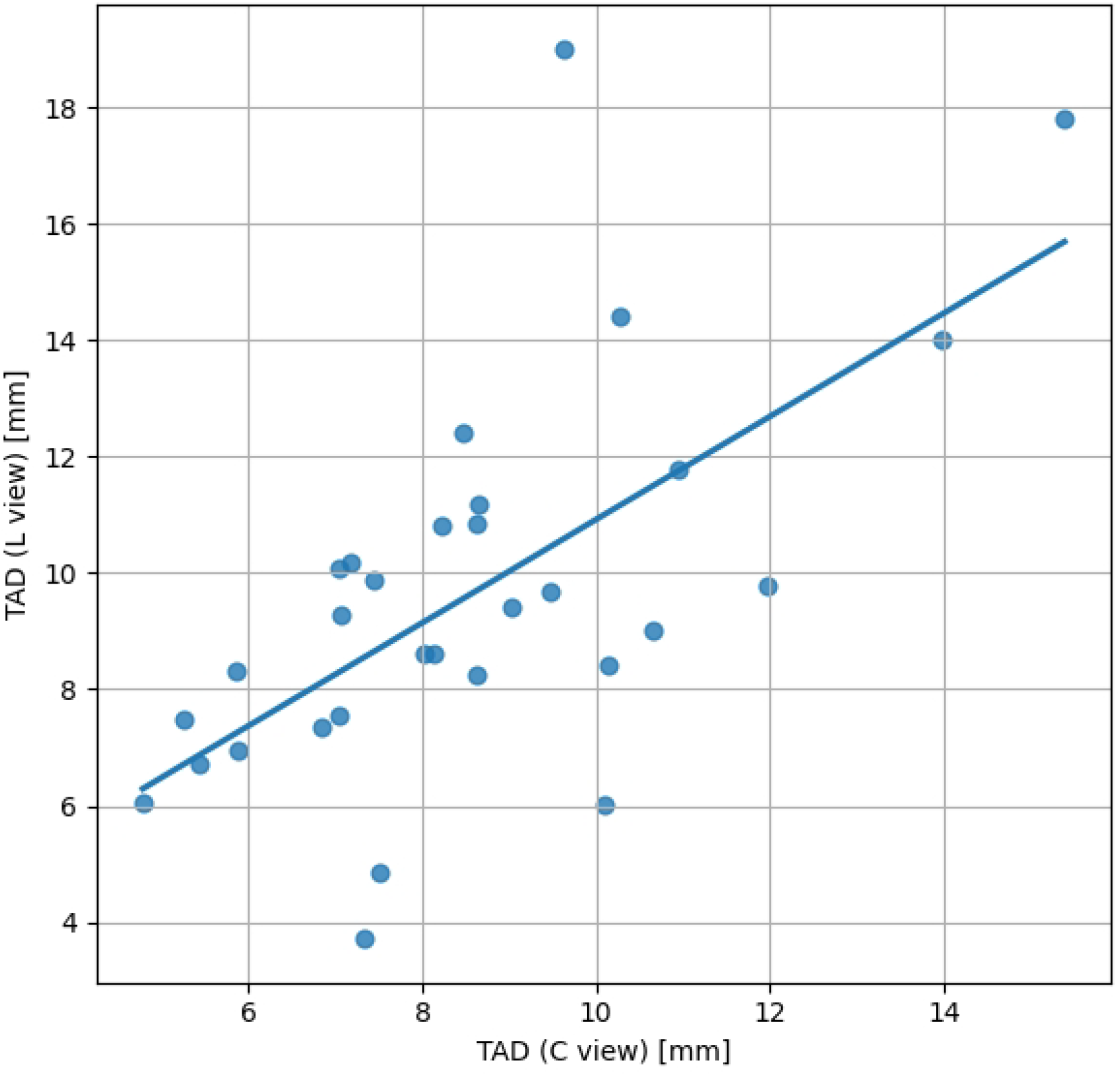
Representative postoperative radiographs from the same patient showing the crosstable lateral view (C view, left) and Lauenstein’s I view (L view, right). The solid line indicates the distance from the screw tip to the apex of the femoral head, measured along the femoral neck axis (dotted line), which constitutes a key component in the calculation of the tip apex distance (TAD).

All procedures were conducted in accordance with the ethical standards of the institutional and national research committees and with the 1964 Helsinki Declaration and its later amendments. Ethical review and approval were not required for this retrospective study using anonymized radiographic data, according to the guidelines of the ethics committee of Nippon Medical School.

The radiographic data were accessed for research purposes on March 15, 2025. All data were fully anonymized prior to analysis, and no author had access to information that could identify individual participants during or after data collection.

## Results

A total of 31 patients met the inclusion criteria. The mean TAD was 8.55 mm (SD, 2.40; 95% CI, 7.62–9.48) in the C view and 9.63 mm (SD, 3.34; 95% CI, 8.41–10.84) in the L view. The difference was statistically significant with a mean of 1.06 mm (p = 0.027; 95% confidence interval: 0.13–2.03).

A significant positive correlation was found between TAD values from the two views (r = 0.637, p < 0.001), as illustrated in Figure 2. Bland–Altman analysis demonstrated good agreement, with most differences lying within the limits of agreement (Figure 3). A summary of TAD values and statistical comparisons is provided in Table 1.

**Figure 2.**
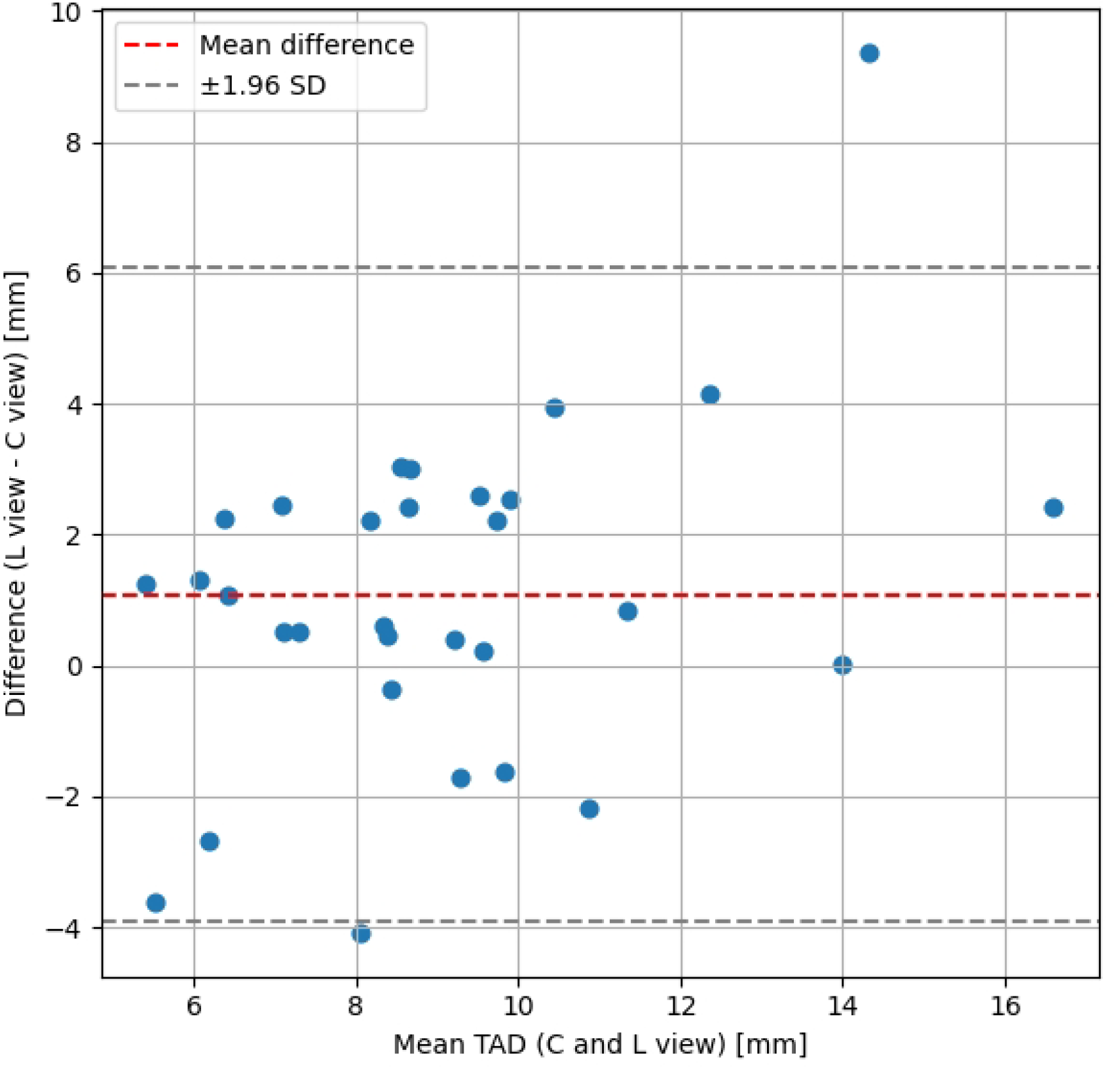
Scatter plot of TAD measured on the cross-table lateral view (C view) versus Lauenstein’s I view (L view). A moderate positive correlation was observed between the two measurements.

**Figure 3.**
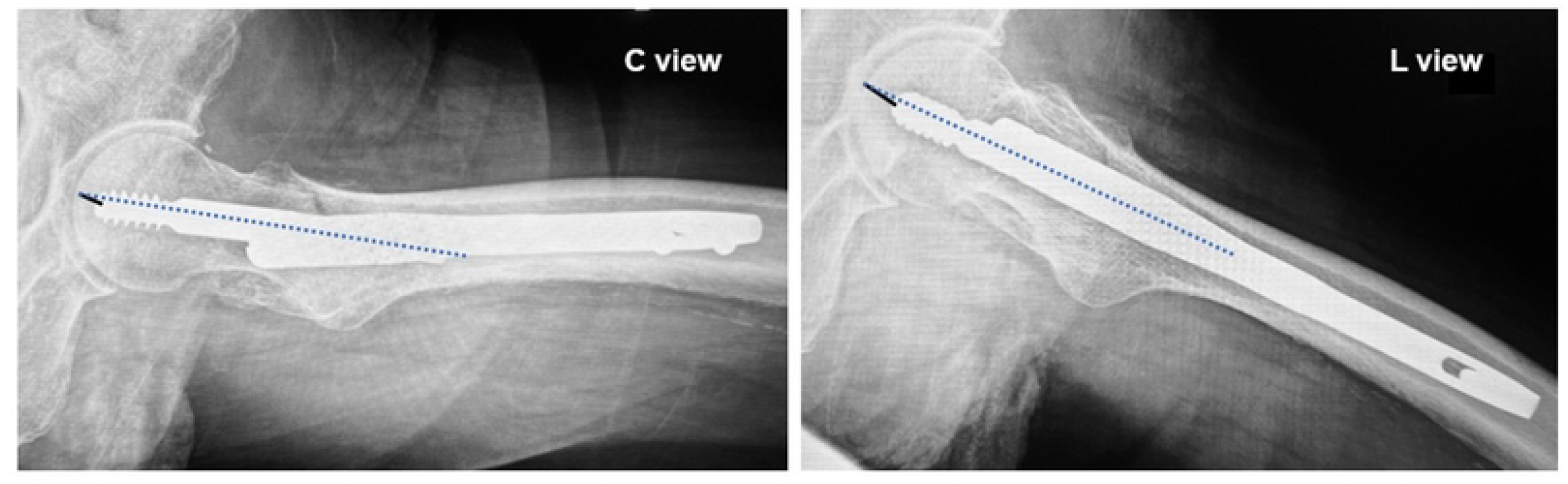
Bland–Altman plot showing agreement between TAD values measured on C view and L view. The mean difference (red dashed line) and the 95% limits of agreement (gray dashed lines, ±1.96 SD) are shown.

**Table 1.** Comparison of tip apex distance (TAD) measurements obtained from the cross-table lateral view (C view) and Lauenstein’s I view (L view). Mean ± standard deviation (SD) and 95% confidence intervals (CI) are shown. A statistically significant difference was found between the two views (p = 0.027).

## Discussion

This study investigated the influence of radiographic projection on TAD measurement by comparing C view and L view following internal fixation of trochanteric femoral fractures. Our findings indicate that although TAD values differed significantly between the two views, the magnitude of the difference was relatively small, and the correlation and agreement were substantial.

Previous studies have emphasized the importance of accurate lateral radiographs in assessing femoral implant positioning and predicting mechanical failure. Nikoloski et al. reported that a rotational error of just 10° in the lateral view could alter TAD by up to 5 mm, underscoring the potential for misclassification. In this context, the L view may serve as a feasible substitute in clinical settings where obtaining an optimal C view is difficult.

Moreover, L view provides broader visualization of the proximal femur, including the greater trochanter and femoral shaft, which may aid in postoperative evaluation. Nevertheless, it is essential to recognize that this study does not assess whether TAD values derived from L view predict cut-out or failure, and therefore its application as a substitute must be interpreted cautiously.

Our findings also align with those of Temmesfeld et al., who demonstrated that C view reliably reflects posterior displacement of the femoral head in Garden type I and II femoral neck fractures, showing a strong correlation (r = 0.86) with 3D-CT measurements (7). Their study supports the notion that C view provides an accurate depiction of femoral head orientation and is robust even when patient positioning is variable. While their work focused on angular posterior tilt and ours on TAD, both studies highlight the importance of lateral radiographs in assessing proximal femoral alignment.

Limitations of this study include the small sample size, retrospective design, and potential selection bias, as only patients who underwent both types of imaging were included. These limitations may affect the generalizability of our findings. Further prospective studies are warranted to validate our findings and to assess whether TAD measurements obtained from L view are predictive of fixation outcomes such as lag screw cut-out.

## Conclusion

Our results suggest that TAD measurements obtained from Lauenstein’s I view, which may offer favorable reproducibility under certain clinical conditions, are generally consistent with those from the cross-table lateral view. These findings indicate that L view could be considered a potential alternative when C view is difficult to obtain. However, whether TAD values measured on L view can predict fixation outcomes, such as lag screw cut-out, remains unclear and warrants further investigation in future prospective studies.

## Data Availability

All relevant data are within the manuscript. Additional datasets generated and analyzed during the current study are available from the corresponding author upon reasonable request.

